# Data model to predict prevalence of COVID-19 in Pakistan

**DOI:** 10.1101/2020.04.06.20055244

**Authors:** Muhammad Qasim, Waqas Ahmad, Shenghuan Zhang, Muhammad Yasir, Muhammad Azhar

## Abstract

Coronavirus disease 2019 (COVID19) has spread to 181 countries and regions and leaving behind 1,133,788 confirmed cases and 62784 deaths worldwide. Countries with lower health services and facilities like Pakistan are on great risk. Pakistan so far has 3,277 confirmed cases and 50 reported deaths due to COVID19. Various mathematical models had presented to predict the global and regional size of pandemic. However, all those models have certain limitation due to their dependence on different variables and analyses are subject to potential bias. As each country has its own dimension therefore country specific model are required to develop accurate estimate. Here we present a data model to predict the size of COVID19 in Pakistan. In this mathematical data model, we used the time sequence mean weighting (TSMW) to estimate the expected future number of COVID19 cases in Pakistan until 29^th^ April 2020. For future expected numbers of cases for long terms the data collection have to be maintained in real time.

## Specification Tables

|  |  |
| --- | --- |
| **Subject** | Infectious Diseases |
| **Specific subject area** | Time sequence mean weighting mathematical model applied to forecast the prevalence and incidence of COVID-2019 in Pakistan |
| **Type of data** | Graphs, Figure, Excel File |
| **How data were acquired** | Data was extracted from http://covid.gov.pk/ |
| **Data format** | Data was in in raw format and have been arranged and analysed in Matlab. An Excel file with data has been uploaded. |
| **Parameters for data collection** | We collected daily reported cases and deaths for COVID19 in Pakistan |
| **Description of data collection** | The daily prevalence data of COVID-2019 from March 10, 2020 to April 1st, 2020 was collected from the official website of COVID-19 Health Advisory Platform by Ministry of National Health Services Regulations and Coordination, Pakistan http://covid.gov.pk/. The Matlab was used to build associated mat files and data was analysed by time sequence mean weighting mathematical model to evaluate the daily reported cases and number of deaths due to COVID-19 in Pakistan. We have calculated the TSMW ($\eta$) and used the model equation $y_n =$ |
| | $(y_{n-1} - d_{n-1})\left(\frac{1}{\eta} - 1\right) - R_{n-1}$ to predict the future state of the given sample space. |
| **Data source location** | University of Otago. Dunedin, New Zealand |
| **Data accessibility** | Raw data and its analysis can be retrieved from the https://github.com/qasimattock/COVID-19-Pakistan.git |

### Value of the Data

- This data will be helpful for Pakistan health authorities to prepare themselves to contain COVID-19.
- As Pakistan is very low in the Global Health Security (GHS) Index therefore they need to utilised their resources in targeted ways and this data will be important for them to take their strategic decisions to mitigate the COVID-19 pandemic in their country.
- Further our model can also use for monitoring and evaluation purpose of their own gown efforts and precautionary measures.

### Data description

The daily prevalence data of COVID-2019 from March 10, 2020 to April 1^st^, 2020 was collected from the official website of COVID-19 Health Advisory Platform by Ministry of National Health Services Regulations and Coordination, Pakistan http://covid.gov.pk/. ^1^ The Matlab was used to build associated mat files and data was analysed by the time sequence mean weighting (TSMW) to predict the future state of the confirmed cases of COVID19 and deaths in Pakistan until 29^th^ April, 2020 (Figure 1). The analysis predicted the expected count of patients 77,905 with minimum 8285 confirmed cases of COVID-19 while expected deaths count of 1382 with lower limit of 45 until 29^th^ April 2020. Currently COVID19 has spread to 181 countries and regions, infected 1,133,788 individuals, and caused 62784 deaths worldwide.^2^ Countries with lower health services and facilities like Pakistan are on great risk.^3^ Pakistan so far has 3,277 confirmed cases and 50 reported deaths due to COVID19.

**Figure:**
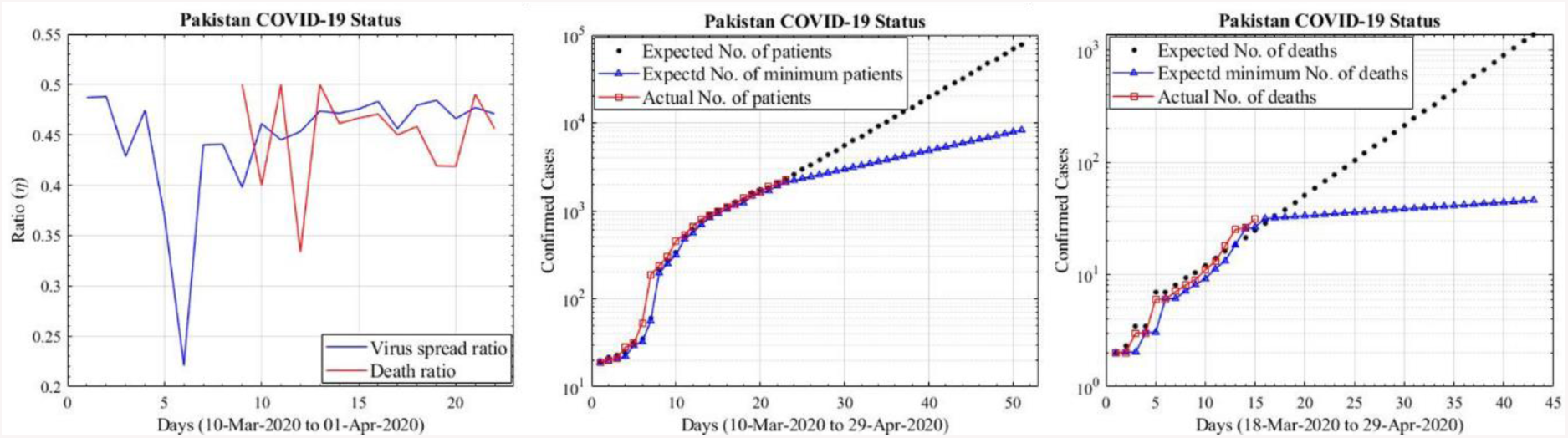
Spread ratio (η) for COVID-19 cases, expected number of cases and expected number of deaths in Pakistan.

### Experimental Design, Materials and Methods

To derived this model, Let *y*_*i*_ represents the number patients reported at *i*^*th*^ day then we define the ratio *x*_*i*_ as

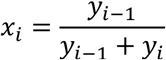

Where, *i* − 1 represents the previous day and the current day is denoted by *i*. The *x*_*i*_ is calculated for all *ϵ* 1, …, *n*, where *n* is the total number of days for which estimation is performed. The mean of all *x*_*i*_ is calculated as ^4^

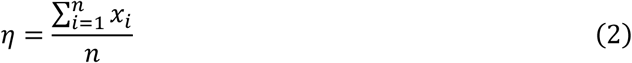

Let *d*_*i*_ represents the total deaths and *R*_*i*_ is the count of patients recovered until *i* days. Then the expected number of patients at day 1 was modelled as

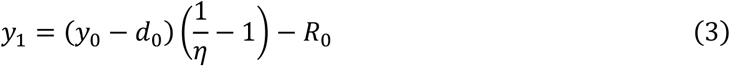

We have assumed the dead patients were eliminated from community without any further virus spreading capability. Similarly, the recovered patients were subtracted from the total number of patients. Extending the Eq. [3] for *y*_2_, …, *y*_*n*_ as

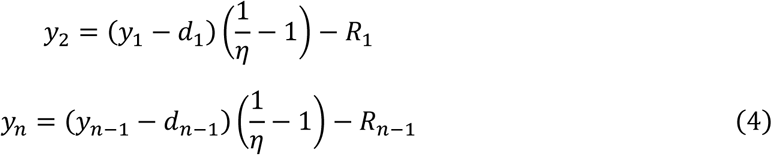

Let *P* be the total population of the sample space then the spread will grow until difference between *P* and *y*_*n*_ is a positive integer defined as

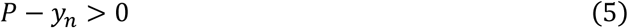

When, *P* − *y*_*n*_ ≤ 0, *n* is the expected number of days at which the whole population will be impacted. The proposed model is different from standard normal distribution as it continuously trains itself from the existing data to estimate the future state. Therefore, it can apply for future real time data.

### Lower bound

Let be the lower bound estimated from *x*_*i*_ as

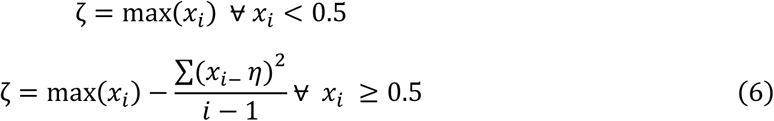

We use *η* = : ζ in Eq. [4] to find the lower bound on the estimated number of patients. The convergence of lower bound remains very close to the actual data set until the same situation prevails and subjected to change with new precautionary measures to contain pandemic.

Our models curves for COVID-19 prevalence and their future trend based on real time data, when new data will come it will further validate our model. If Pakistan does not act on complete lockdown and strict precautionary measures due to its week health system its whole population will be on risk.^3^ Our data shows that it will take roughly 90 days (1-July-2020) to affect the 95% population under the constraint that there is no isolation policy implemented among the population.

## Data Availability

Data was extracted from http://covid.gov.pk/
Raw data and its analysis can be retrieved from the https://github.com/qasimattock/COVID-19-Pakistan.git

## Appendix A

### Supplementary data

Supplementary data to this article can be found online at https://github.com/qasimattock/COVID-19-Pakistan.git

